# Cortico-Spinal Intermittent Theta Burst Stimulation Propelling Sensorimotor Function Recovery in Complete Spinal Cord Injury: Randomized Control Trial Protocol

**DOI:** 10.1101/2024.04.13.24305754

**Authors:** Deeksha Patel, Rohit Banerjee, Kamran Farooque, Deepak Gupta, Bhavuk Garg, Nand Kumar, KP Kocchar, Suman Jain

**Author notes:** **Corresponding author-** Suman Jain (Professor), **Email Addresses:** (Suman Jain).

## Abstract

**Background:** Intermittent theta burst stimulation (iTBS) is a non-invasive stimulation technique to induce neuronal and synaptic plasticity. The induced cortical plasticity is imperative in the recovery of motor and sensory functions. Spinal cord injury (SCI) causes damage to neurons and results in sensorimotor dysfunction. The effect of iTBS on recovery of motor and sensory dysfunction in complete SCI (cSCI) is still elusive.

**AIM:** This study aims to assess the effect of iTBS on cortico-spinal tract integrity, plasticity, and regaining of motor and sensory function in cSCI patients. The rationale behind using an iTBS protocol is to modify and augment the communication between spared neurons of the cortico-spinal tract and strengthen the synaptic transmission, which will improve motor function in underlying muscles.

**Method:** A total of 48 patients will be recruited and randomly divided into placebo and real stimulation groups. iTBS along with a rehabilitation program will be administered to the placebo and real stimulation groups. Follow-up will be done at 1 month, 2 months, and 3 months after the intervention.

**Result:** The outcome of the study will be defined by electrophysiological parameters elicited by single and paired-pulse stimulation, ASIA score, pain, activities of daily life, quality of life, anxiety, depression, and biomarkers related to SCI. The results of this study will uncover the effectiveness of iTBS stimulation on (i) recovery of motor and sensory function in cSCI (ii) excitability of cortico-spinal tract (iii) neurological recovery and modulation of pain (iv) cortical reorganization after injury.

**Conclusion:** Intermittent theta-burst stimulation (iTBS) in conjunction with an individualized rehabilitation program may serve as an integrated strategy to rejuvenate locomotor abilities and improve the overall quality of life for people with complete spinal cord injuries (SCI).

**Key Points:**

1. iTBS is a novel neurostimulation technique aimed to restore sensorimotor function after a complete SCI
2. The primary objective of the trial is to evaluate the efficacy of iTBS in promoting sensorimotor function recovery
3. Assessment of the potential impact of iTBS on SCI rehabilitation
4. Understand the underlying mechanism of excitatory-inhibitory circuits associated with SCI
5. Unlock the importance of neuronal plasticity in regaining mobility

## Introduction

Spinal cord injury (SCI) is a neurological condition that causes progressive neurodegeneration. The major symptoms of SCI are paralysis, paraesthesia, pain, spasticity, bladder, bowel, and sexual dysfunction. It largely affects the young age population (21-49 years), thus the psychological impact on a healthy individual to adapt to a paraplegic or quadriplegic condition in their early life is devastating. SCI causes damage to neurons, nerves, and other surrounding cells that send and receive signals from different body parts to the brain via the spinal cord and vice versa(1). Spinal cord injury can be traumatic or non-traumatic. Traumatic spinal cord injury (SCI) is a great challenge for therapeutic management considering its high morbidity. There is complete or incomplete loss of locomotor, sensory, and autonomic functions depending on the severity and level of the lesion(2). The consequences of injury are not just a break in communication between neurons, but a cascade of events that sets up a vicious cycle and leads to widespread neuronal degeneration, cell death, and the formation of glial scars(3). Cellular components such as proteins, phospholipids, neurotransmitters, and metabolites derived from SC neurons and glial cells diffuse from the injury site into the cerebrospinal Fluid (CSF) and blood(4). These biomarkers could serve as good diagnostic markers to predict the severity of the injury. To regain functional connectivity, and attenuate gliosis and secondary injury, activity-dependent strategies have been proposed to be quite effective. The current treatment modalities for SCI include surgery, pain management, and rehabilitation. However, functional recovery by restoring connections of the corticospinal tract after a complete spinal cord injury is challenging.

High-frequency repetitive transcranial magnetic stimulation has decreased neuropathic pain and spasticity in incomplete spinal cord-injured patients(5)(6). A newer form of patterned TMS, intermittent theta burst stimulation (iTBS) is now being studied for the improvement of symptoms in incomplete spinal cord injury(7). There is very limited literature available, showing the effect of either cortical iTBS or trans-spinal TMS in combination with a rehabilitation program to promote repair, regeneration, and recovery in cSCI patients The present study aims to determine the functional outcomes of administering iTBS at the cortex as well as at spinal cord along with intensive rehabilitation program in cSCI patients.

### Objectives

1. To establish an effective site for intermittent theta burst stimulation (iTBS) in complete SCI patients: Motor cortex/ Spinal cord / Motor cortex + Spinal cord
2. To evaluate the efficacy of iTBS along with rehabilitation programs on –
  a. Recovery of sensory and locomotor functions
  b. Cortical excitability and plasticity
  c. Biomarkers associated with spinal cord injury
  d. Pain, anxiety, depression, and quality of life scores

## Material & Methods

### Ethical Approval

Ethical approval has been obtained from the Institutional Ethical Committee of All India Institute of Medical Sciences, Delhi (Project Ref. Id: IECPG/551/7/2022). The study is registered in Clinical Trials Registry-India (CTRI) with reference number CTRI/2022/11/047038. Informed, voluntary, and written consent will be taken and the participants will be given a choice to withdraw from the study at any given point in time. They will be duly informed of the duration of the study along with any potential risks or side effects associated with TMS intervention.

All the experiments will be carried out in the Brain Stimulation and Neuromodulation Laboratory, Department of Physiology, and CARE, dept of Psychiatry, AIIMS, New Delhi.

### Study Design

A double-blinded, prospective, randomized, placebo-controlled study will be conducted. The computer-generated sequence will be sealed in sequentially numbered envelopes, which will be opened after the patient is enrolled in the study. Both the patient and investigator will be blinded to the intervention. An experienced therapist will perform the intervention protocol. The study will follow the CONSORT rule (Figure 1).

**Figure 1.**
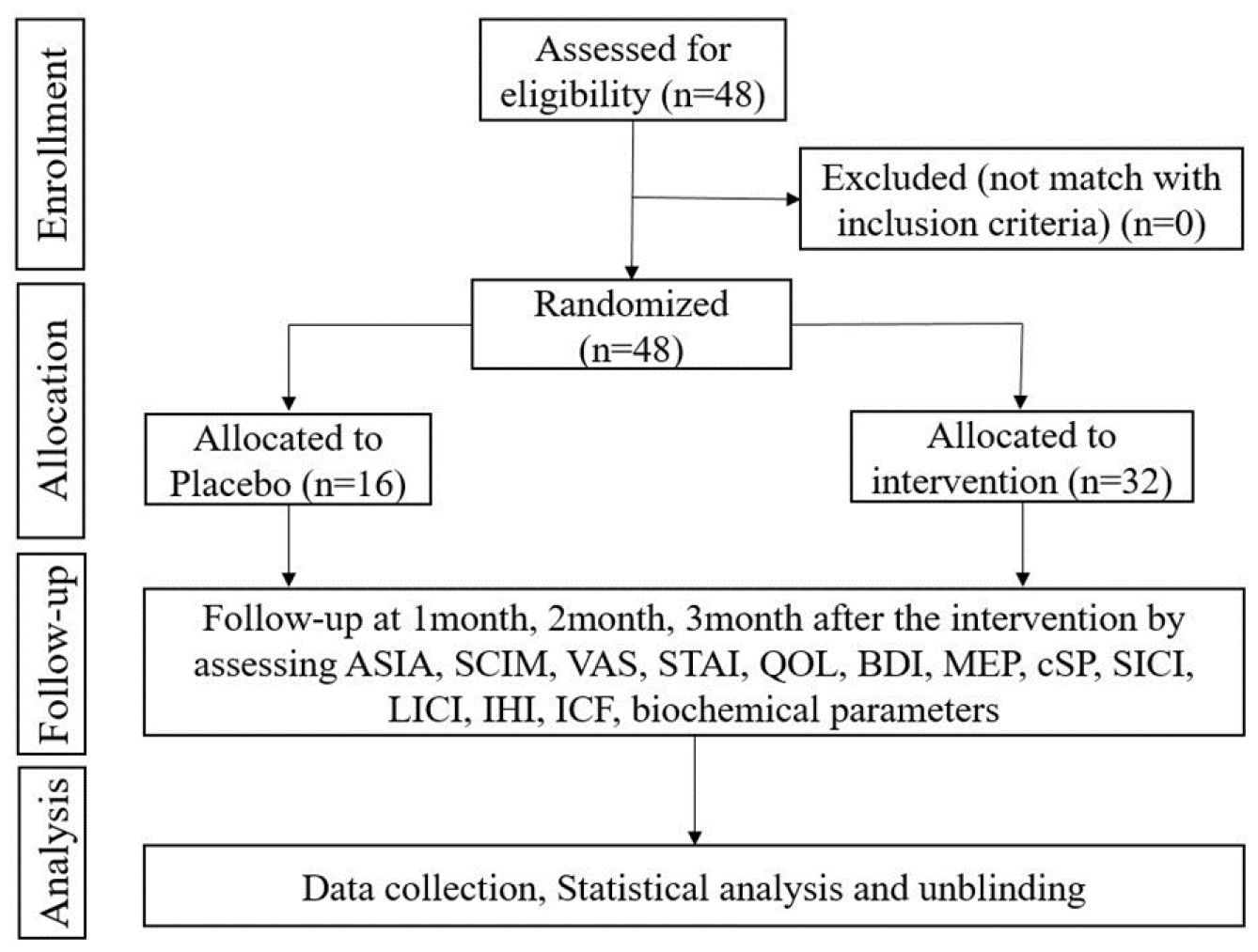
CONSORT diagram. ASIA: American Spinal Cord Injury Association; SCIM: Spinal Cord Injury Independence Measure; VAS: Visual Analogue Scale; STAI: State-trait Anxiety Inventory; QOL: Quality of Life; BDI: Beck depression inventory; MEP: Motor evoked Potential; cSP: Cortical silent period; SICI: Short interval intracortical inhibition; LICI: Long interval intracortical inhibition; IHI: Interhemispheric inhibition; ICF: Intracortical facilitation

### Duration of study

The recruitment, intervention, and follow-up phase will be of 2 years, followed by 6 months of data analysis. The recruitment of patients will be started in September 2022 and the trial will be completed by March 2025.

### Inclusion-Exclusion Criteria

Adults in the age group of 18 to 60 years with Thoracolumbar Spinal cord Injury having complete motor loss below the level of lesion with ASIA score A will be screened and recruited in the study. Patients with osteoporotic Fracture, having a history of neurological or orthopedic disease related to the spinal cord, head injury, ferromagnetic metallic implants close to the target stimulation area, pacemaker, cognitive impairment, pregnancy, history of seizures and acute eczema/ bed shore will be excluded from the study.

### Confidentiality

The personal privacy of patients will be protected. The results of this study will not disclose any identifying and personal information of the patient without his/her permission.

## Experimental Groups

A total of 48 patients will be randomized into 6 groups using computer-generated random numbers:

1. Group A: Control
2. Group A: Sham (iTBS with placebo coil on motor cortex)
3. Group B: High-frequency rTMS on motor cortex
4. Group C: iTBS on motor cortex
5. Group D: iTBS on the spinal cord
6. Group E: iTBS on the motor cortex and spinal cord

### Intervention

The rTMS/ iTBS will be applied using Neurosoft - Neuro-MS 5 (Neurosoft Ltd®, Ivanovo, Russia), a commercially available transcranial magnetic stimulator equipped with an angulated figure-eight-shaped coil and circular coil will be used for stimulation.

The intervention will be applied over the lower-limb motor area localized in M1 (to stimulate both lower limbs), with the handle of the coil parallel to the interhemispheric midline (pointing occipitally) based on the vertex position of the International 10-20 system of EEG(8,9). For spine stimulation, the circular coil will be placed over the site of injury at the spinal cord.

Standard protocol of iTBS will be used; 3pulse bursts at 50 Hz, repeated at 5 Hz, 2-s train repeated every 10 s for 20 repetitions, for a total of 600 pulses (7). 90% resting motor threshold will be used as the iTBS intensity. rTMS consists of 1600 pulses at 20Hz frequency. 2-s train of rTMS will be repeated every 30 sec for 20 minutes(10). A 90% resting motor threshold will be used as rTMS intensity.

2 weeks postoperatively, rTMS or iTBS will be administered in 2 sessions/day for 5 days, with a total of 10 sessions. The protocol and number of sessions for the sham stimulation group will be the same as that for the actual stimulation group, except the coil will be a placebo. (Figure 2)

**Figure 2.**
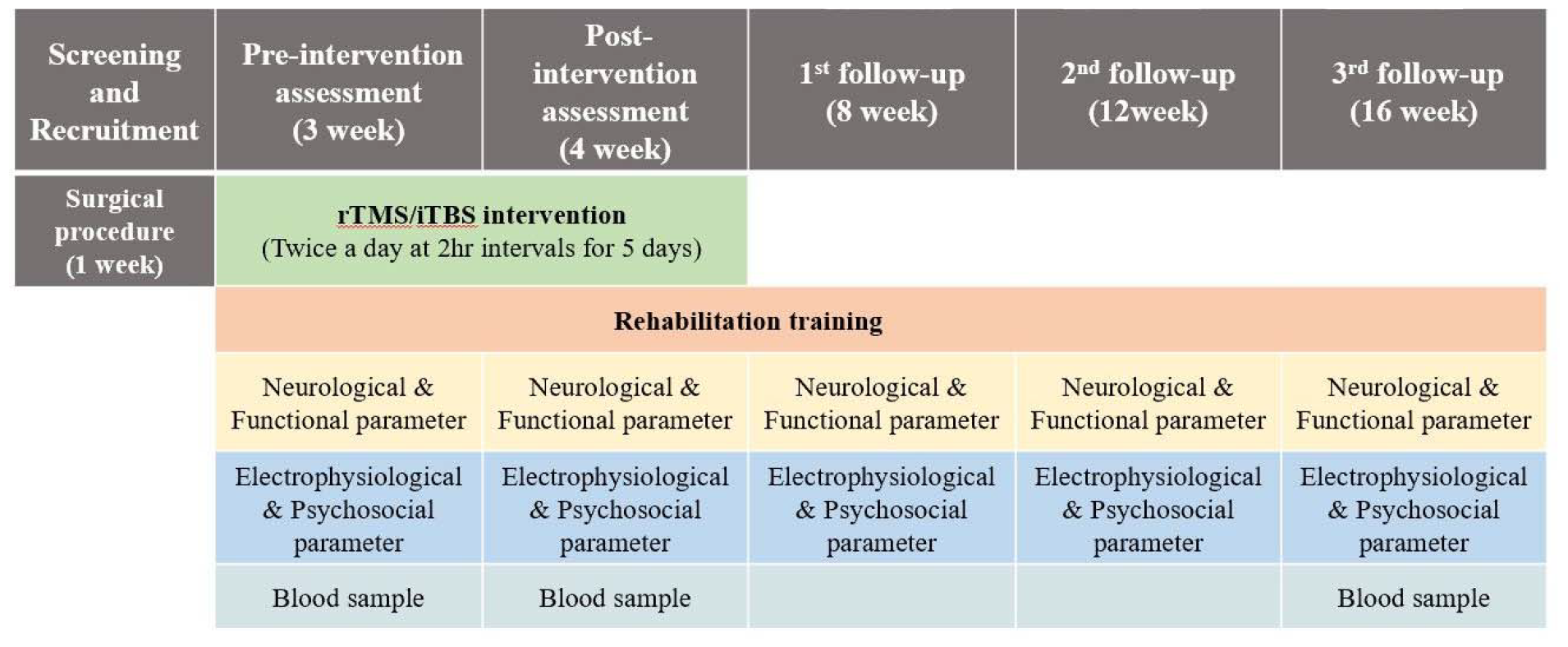
Timeline-. All the intervention and recording paradigms are organized as a timeline

### Methodology

1. Patients will be recruited from J P N A Trauma Centre, AIIMS Delhi after the vertebral injury has been stabilized. At the first visit, the baseline parameters of the patient will be assessed.
2. During an intervention, the patient will be allocated to one of the interventional groups as per randomization. Motor cortex stimulation will be followed by spinal cord stimulation for 10 sessions twice a day for 5 consecutive days.
3. For cortical TMS intervention, firstly the Resting motor threshold (RMT) will be determined using MEP recorded from Abductor Pollicis Brevis (APB). The figure of eight coils will be used over motor hotspots of APB The cortical stimulation intensity used to generate MEP in target muscle with amplitude greater than 50μV in 5 out of 10 consecutive trials will be considered as RMT. The stimulation intensity for iTBS or rTMS will be kept at 90% of RMT and the coil will be placed at the leg area of the M1 area. In the Sham stimulation group, the protocol will remain the same, but the stimulation coil will be placebo.
4. For spinal TMS intervention, the patient will be lying in a prone position. The figure of eight coil will be held parallel over the vertebral column corresponding to an injured area of the spinal cord. The stimulation protocol will remain similar to cortical stimulation.
5. A trained physiotherapist will be assigned to provide personalized rehabilitation training for 5 days (Figure 3). Thereafter the patient will be guided to perform all the exercises at home and maintain a logbook, share their videos of performance weekly for assessment by the physiotherapist.
6. Blood samples will be collected before intervention, immediately after intervention, and after 3 months of intervention.

**Figure 3.**
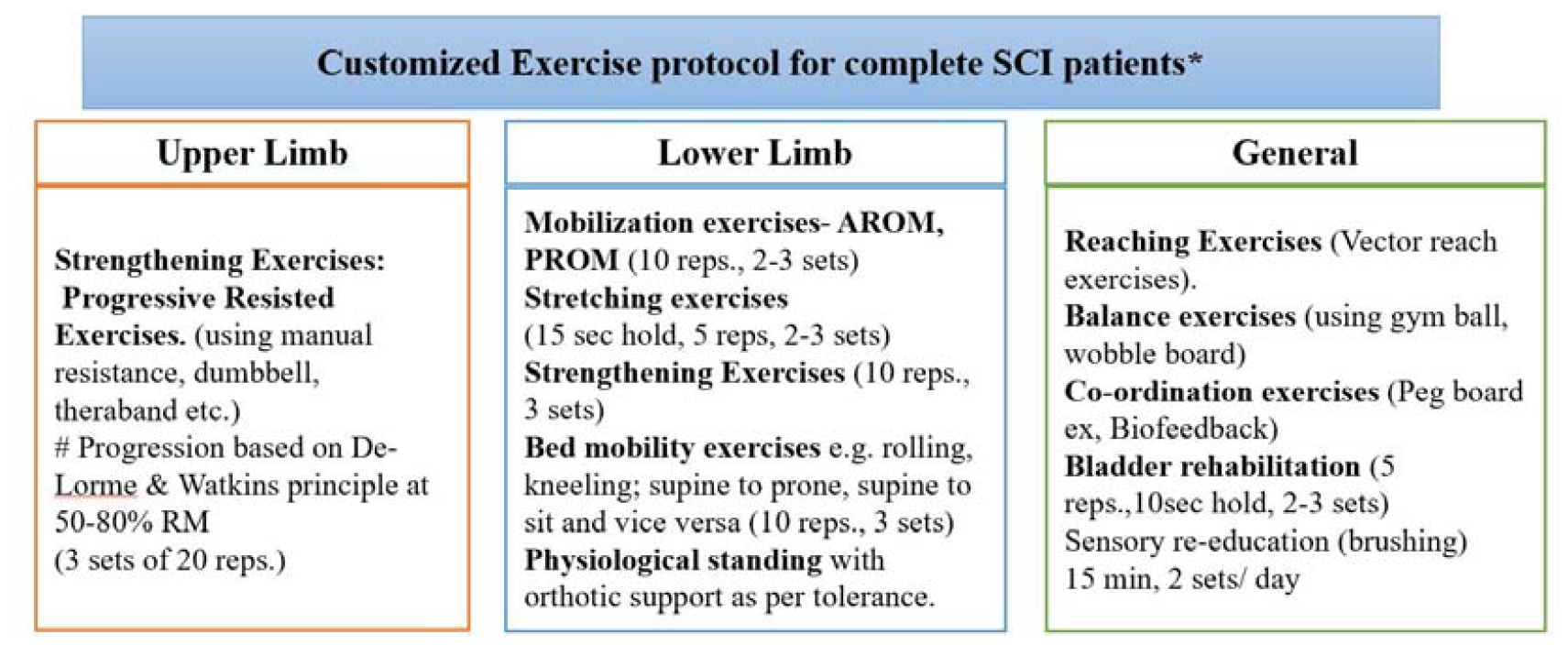
Outline of physical rehabilitation regime for spinal cord injury patients. 9. ^*^Exercise prescription and progression will be based on the individual’s performance and tolerance to baseline exercises.

### Post-intervention care

If a patient experiences an adverse event during the trial, the principal investigator will provide treatment and corresponding financial compensation. Patients who are enrolled in the placebo will also receive the conventional rehabilitation program during the study.

## Results

The outcome will be measured at 5-time points: baseline, immediate post-intervention, 1^st^ month, 2^nd^ month and 3^rd^ month of follow-ups. All the assessments will be recorded by the blinded primary investigator to maintain the discrepancy during the intervention (Table 1).

**Table 1.**
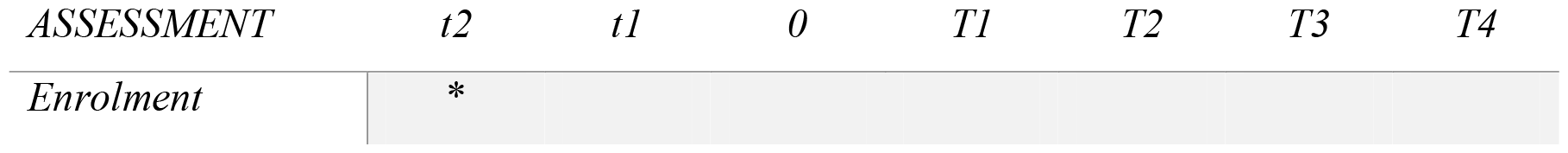
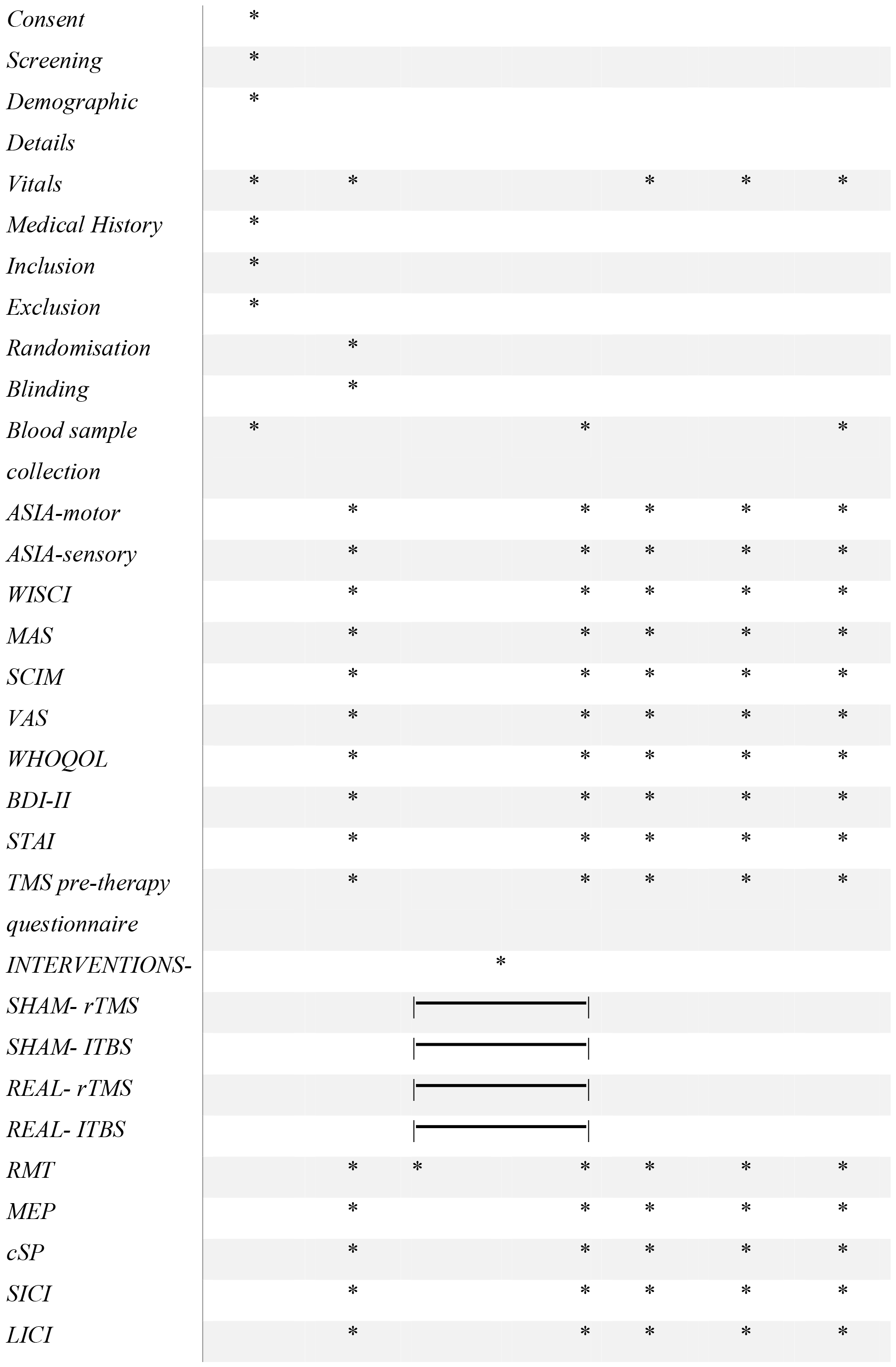
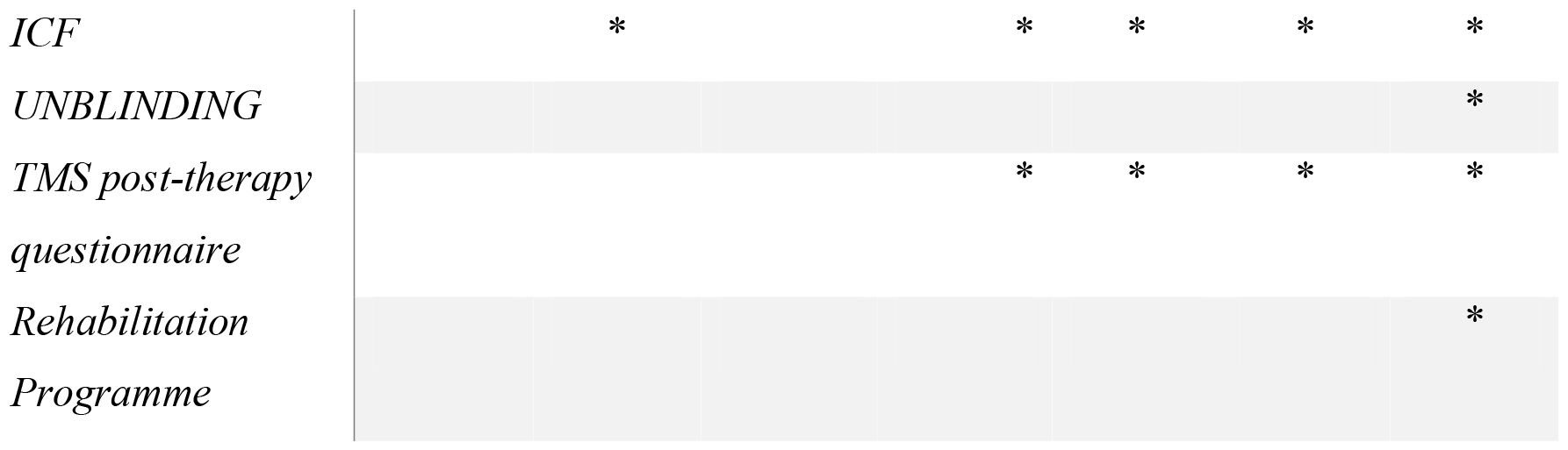
Protocol for intervention trial and outcomes measure will be recorded at different time intervals. −t2: Screening before surgery; −t1: Parameters assessment day before intervention; 0: intervention start for consecutive 5 days; T1: Parameters assessed after completion of intervention; T2: follow-up at 1^st^ month; T3: follow-up at 2^nd^ month; T4: follow-up at 3^rd^ month.

1) **Neurological:** The primary outcome will measure the severity of injury with the American Spinal Injury Association (ASIA) Scoring System. The ASIA impairment score (AIS) ranges from complete loss of sensation and movement (AIS = A) to normal neurological function (AIS = E). The ASIA motor score uses a test of the strength of ten key muscles on each side of the body (e.g. elbow flexors, wrist extensors, hip flexors, quadriceps, dorsi flexors). The score ranges from 0 (no contraction) to 5 (normal resistance) through a full range of motion. A total possible score of 50 for the upper extremities (UE) and 50 for the lower extremities (LE) may be obtained (11). The ASIA sensory score involves pinprick and light touch sensation at key points representing each dermatome of the body, scored on a three-point scale (0, 1, and 2). Scores are summed to give a total possible score of 224, where a higher score indicates better sensation than a lower score.
2) **Functional**
  a. Walking Index for Spinal Cord Injury II (WISCI-II) is a 0–20 level scale, that evaluates the walking activity of a patient based on physical assistance, the need for braces/walker, and other adaptive devices. The levels on the scale are scored from 0 (patient unable to walk) to 20 (patient walking without braces and/or adaptive devices and without any physical assistance for at least 10 m)(12).
  b. Spinal Cord Independence Measure-III (SCIM-III) includes 19 tasks organized in 3 subclasses based on the patient’s general activity: self-care (score 0-20), respiration and sphincter management (score 0-40), and mobility (score 0-40). The overall scores range from 0-100, where a 0 score defines a total dependence of the patient on the caregiver and a score of 100 indicates the complete independence(13).
3) **Electrophysiological:**
  a. **Single-pulse TMS:** A single pulse of TMS at minimum stimulus intensity will be delivered at the motor cortex that elicits a motor-evoked potential (MEP) of ≥ 50μV at least 5 out of 10 trials in the target muscle and will be recorded as RMT(14). For recording the inhibitory activity, a single TMS is delivered to produce an interruption in ongoing EMG activity during a tonic contraction followed by the reoccurring EMG activity. The duration of silencing of EMG in response to TMS will be measured as cSP (15). The recruitment curve plotted between the **TMS** intensity (%) and motor-evoked potential (MEP) sizes.
  b. **Paired-pulse TMS:** Two TMS pulses, conditioning stimulus (CS) and test stimulus (TS) delivered at specific interstimulus intervals result in either facilitation or inhibition. In SICI, MEP elicited by TS is inhibited when preceded by a CS at ~1-5ms intervals. In LICI, the interstimulus interval is kept between 50-200ms, whereas, in ICF, the interval is ~10-30ms (15).
4) **Psychosocial:**
  a. Visual Analogue scale (VAS) measures the perceived intensity of pain on a self-explained scale of 0 to 10, where 0 (no pain) to 10 (severe pain)(16).
  b. Beck depression inventory II is a 21-question multiple-choice self-report inventory to measure the presence and severity of depression. The cut-scores with 0-9 indicating normal, 10-19 indicating mild depression, 20-30 indicating moderate depression, and 31-63 indicating severe depression(17).
  c. State-Trait Anxiety Inventory is a 40-item self-report scale, commonly used to measure anxiety (18).
  d. The WHO-Quality of Life-BREF Scale is developed in the context of the four domains defining the QOL: physical, psychological, social, and environmental. The higher the QOL score the higher the life satisfaction(19).
5) **Biochemical Quantification:** The disruption of blood spinal cord barrier (BSCB) and secondary damage following the injury releases several chemokines, growth factors, and neurotransmitters in CSF and systemic circulation which could help predict the severity of injury. The released components such as Myelin Basic Protein (MBP), Interleukins, Phosphorylated neurofilaments, BDNF, Fas-ligand, GABA, and Glutamate may serve as potential biomarkers for remyelination, inflammation, neuronal survival, and excitatory or inhibitory neurotransmitters after SCI (20)(21)(22)(23). Mass spectroscopy and enzyme linked immunosorbent assays will be performed to quantify these biomarkers in plasma and serum samples.

## Statistical Analysis

Data will be presented as Mean and standard deviation (SD). The intention-to-treat (ITT) analysis will be the primary analysis. Baseline observation will be carried forward for subjects with missing values. Unless otherwise stated, ITT data will be presented throughout. Per-protocol analysis (all compliant participants) will also be conducted. To see the difference in groups ANOVA analysis followed by multiple comparison tests (Post hoc analysis) will be used. To see the changes at different time points from the baseline, paired t-test or repeated measure ANOVA will be applied. To establish an association between groups and qualitative variables, the Chi-square/fisher exact test will be used. The value of p less than .05 will be considered statistically significant. P value < 0.05 at (95% CI) will be considered statistically significant. Statistical analysis will be done using STATA 14.0 (STATA Corp, Houston, TX, USA) software.

## Discussion

SCI significantly impairs the cortico-spinal integrity and afferent-efferent input/output circuitry. Theoretically, the pyramidal tract is the primary neural pathway that links the cortex and spinal cord to facilitate the movement of distal extremities. The primary purpose of any treatment is to reconstruct the neural circuit immediately after SCI for functional sensory-motor recovery. To facilitate neural circuit reconstruction, it is required to stimulate nerve cell sprouting and regeneration as well as increase the strength of the existing neuronal connections. Previous research demonstrated that individuals with incomplete SCI can benefit from locomotor training to enhance their motor skills. Rehabilitation programs may use learning and relearning mechanisms, uncovering a previously inactive synapse, and forming a new synapse(24). However, the reconstruction of the damaged neural circuit is quite difficult with locomotor training alone. Studies have shown that the functional effects of exercise along with transcranial magnetic stimulation, can activate spared neural pathways and enhance the possibility of neural reconstruction(25,26).

Transcranial magnetic stimulation (TMS) induces electrical currents in underlying cortical areas, depolarizes neurons, and generates an action potential that modulates the activity of spinal motor neurons and target muscles via the corticospinal tract. Repetitive transcranial magnetic stimulation as well as iTBS is an intervention in various psychiatric and pain conditions. In SCI few studies suggest improvement in locomotor function, spasticity, and pain in incomplete patients(6). Although, the functional mechanism of rTMS or iTBS on sensorimotor recovery in SCI patients is not fully understood, but thought to induce synaptic plasticity via LTP/LTD-like effects (27), thereby can promote functional recovery in SCI patients.

Additionally, spinal cord stimulation can modulate the activity of the local central pattern motor generators, which promote synaptic strengthening (28). In rat models of complete and incomplete SCI a significant attenuation of glial scaring, lesion volume, neurotransmitter imbalance, muscle atrophy, and facilitation of neuronal survival, axonal regeneration, and myogenesis has been shown following whole-body magnetic field exposure(29,30). Therefore, we propose that spinal cord stimulation along with motor cortical stimulation would attenuate secondary damage and promote regeneration even in complete SCI patients, leading to long-term functional recovery.

Various single and paired-pulse paradigms of TMS have been used for the assessment of cortical excitability, plasticity, integrity of the corticospinal tract, and excitatory-inhibitory (E-I) neural circuitry of the motor cortex. We shall use all paired-pulse paradigms (SICI, ICF, and LICI) to objectively assess E-I neural circuitry and single-pulse paradigms (RMT, MEP, recruitment curves, and cSP) for unraveling the excitability of corticospinal circuitry and motor units.

## Conclusion

An intensive individualized rehabilitation regime coupled with iTBS could be a holistic management strategy that can restore locomotor function and quality of life in complete SCI patients. TMS is also a promising tool to evaluate the cortical plasticity and excitability in SCI and thereby understand the mechanism of action of the proposed intervention. characterize the effective connectivity of neural circuits and mechanisms regulating the balance between inhibition and facilitation within the corticospinal pathway.

## Data Availability

All data produced in the present study are available upon reasonable request to the authors.Though this is an protocol so doesn't contain any data.

## Acknowledgment

We want to acknowledge the Department of Biotechnology (DBT) for providing us with funding. We also want to acknowledge Mr. Sanjeev and Mr. Praveen for this technical support during standardization.

